# Arm Angle Moderates the Association Between Fastball Usage and Elbow/Forearm Injury in MLB Pitchers

**DOI:** 10.64898/2026.08.29.26361727

**Authors:** Connor Richards, D. Taylor La Salle, Oscar Vila Dieguez, Samuel R Ward

## Abstract

**Background:** Newly available arm angle data offers a new dimension to understand rising rates of arm injury in MLB pitchers.

**Purpose:** To evaluate the relationship between arm angle, pitch characteristics, and elbow and forearm injury in MLB pitchers.

**Study Design:** Retrospective cohort study; Level of evidence, 3

**Methods:** Statcast data from 2020 to 2025 and MLB injured list (IL) data were used to evaluate arm angle and pitch characteristics in relation to elbow and forearm injuries. Results are presented with and without requirements on prior season workload and for same-season and next-season injury incidence. A generalized additive model (GAM) was used to capture non-linear dependence and interactions between selected features and injury incidence to the elbow or forearm. Average marginal effect (AME) odds ratios are reported for main effect terms.

**Results:** N = 3,812 pitcher-seasons were included. 29% pitchers who underwent UCLR did so in the same season as a forearm injury (*t_mean_* = 44, *t_median_* = 27 days to surgery). Arm angle, fastball usage, and their interaction were the three most predictive features. Arm angle was positively related to incidence of injury (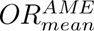 = 1.014), fastball usage was inversely related to incidence of injury (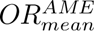 = 0.243), and arm angle moderated the effect fastball usage at high arm angle, where increased usage was no longer protective. Slider velocity (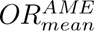 = 1.072), spin rate (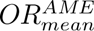 = 1.001), and usage (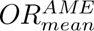 = 2.039) also significantly predicted injury risk. Fastball velocity was not significant in any fit, with 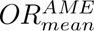 = 0.999 across all fits. Fit-level Nagelkerke *R*^2^ values ranged from .019 to .052.

**Conclusion:** Fastball usage and arm angle, not velocity, predicted elbow and forearm injury risk among MLB pitchers, and arm angle was the single most predictive feature. The heterogeneity of risk factors as a function of arm angle, and the novelty of MLB arm angle data, may explain why fastball usage has been previously underexplored as a risk factor.

## Introduction

Medial ulnar collateral ligament (UCL) injuries are one of the most pressing problems facing Major League Baseball today. From 2011 to 2023, medial UCL injuries accounted for 583,070 cumulative days missed among MLB players, more than any other injury.^27^ The rising rates of upper-extremity injuries, including elbow and forearm injuries, only underscore the importance of understanding injury risk among pitchers.^17,23,27^ Despite this, pitcher risk factors for elbow injury, and their interrelationships, are not fully understood.

The relationships between velocity, spin rate, and injury are well-studied, but some results are in tension with each other, particularly when it comes to pitch usage. Numerous authors have identified a link between pitch velocity and injury.^3,16,19^ Results have linked overall pitch velocity to UCLR,^3^ overall pitch velocity to incidence of elbow injury,^19^ and fastball velocity to UCLR;^16^ slider spin rate has also been identified as a significant predictor of UCLR risk among MLB pitchers.^16^ However, Keller et al. (2016) found no significant association between fastball velocity and UCLR, instead observing a significant effect of fastball usage.^7^ One challenge facing analyses of medial elbow injuries is that UCL reconstruction among MLB pitchers, while altogether too frequent and the largest injury problem facing professional baseball, occurs infrequently relative to the sample size required for the more complex approaches capable of assessing the interaction of these factors.

While authors have historically focused on UCLR, evidence of the importance of forearm injuries and their relationship to UCL injuries in professional pitchers is mounting. Forearm injuries have been identified as an immediate precursor to UCLR, with Hodgins et al. (2018) finding that 19.4% of MLB and Minor League Baseball (MiLB) players sustaining a forearm injury required UCLR within one year.^5^ Likewise, Zaremski et al. (2022) found a significant association between forearm flexor injury and UCLR.^30^ The recent rise in forearm and flexor tendon injuries since 2023^23^ and the aforementioned link to UCL injuries has led to authors like Freehill (2026) calling for increased attention to the forearm, forearm injuries, and the connection to UCL injuries.^18^ This epidemiological overlap has an anatomical basis. The flexor-pronator muscles arise from the medial epicondyle immediately superficial to the anterior bundle of the UCL, and cadaveric work has shown that the deep aponeuroses of the flexor digitorum superficialis and flexor carpi ulnaris, the tendinous septa between them, and the anterior bundle form a single tendinous complex that cannot be separated histologically.^6^ Functionally, these muscles act as secondary dynamic restraints to valgus load, with the flexor carpi ulnaris producing the greatest reduction in valgus angulation and the flexor digitorum superficialis the next greatest in a UCL-deficient elbow.^21^ Because the ligament and the flexor-pronator mass share an origin, a load path, and a stabilizing role, load exceeding the tolerance of one raises the demand on the other. This gives an a priori reason to treat elbow and forearm injuries in pitchers as two presentations of the same medial elbow complex rather than as independent outcomes. This interest in forearm injuries and their link to medial elbow injuries means that forearm injuries among MLB pitchers represent both an important epidemiological signal and additional sample to study arm injuries.

Oeding et al.’s (2024) machine learning approach to predicting elbow injury was able to probe non-linear relationships between pitch-tracking variables because they studied placement on the MLB Injured List (IL), not UCLR.^19^ Where prior analyses were constrained by limited sample size for the strong signal of UCL reconstruction, this approach swapped the low-incidence, high-fidelity target of UCLR for a high-incidence, lower-fidelity target of IL placements. This allowed them to probe interactions between velocity, usage, and spin rate, among others, by greatly increasing the effective sample size.^19^ This approach suggests that non-linear interactions between pitch-level features offer signal for injury risk and that MLB IL data offers the sample sizes required to explore them.

Unlike velocity and spin rate, arm angle has been comparatively underexplored in the literature, and for good reason. While some studies in a lab or simulated game setting evaluated the relationship between arm angle and injury, e.g. Okoroha et al. (2018), these results mostly focus on intrasubject changes in arm angle as predictors of fatigue.^20^ Lipa et al. (2025) examined arm angle as a potential predictor of UCLR across subjects, but their intensive, manual data collection of arm angle from game film limited them to 20 pitches per subject, and they found no significant effect.^11^ The injury literature to date has been limited in this area by the lack of large-scale collection of arm angle data for MLB pitchers.

Following the 2024 MLB season, Statcast added arm angle to the list of pitch-level metrics tracked in all games.^1^ To date, arm angle data has been backfilled to give coverage from 2020 onward, meaning more than five full seasons of pitch-level arm angle data are available. Combined with Oeding’s approach to use IL placements to increase effective sample size, this offers a rich dataset to explore questions raised by Keller, Lipa, and others.

We propose using pitch-level arm angle data, along with velocity, spin rate, and usage, to evaluate their relationship to incidence of elbow and forearm injury from MLB IL data.

## Methods

Pitch-level Statcast data for regular season MLB games from the 2020 through 2025 seasons (July 2020 to October 2025) was accessed via the pybaseball Python package.^9^ UCL reconstruction (UCLR) was confirmed via a public database of pitcher surgeries.^26^ MLB transaction data was obtained using the MLB Stats API.^13^ Data retrieval and visualization were performed using Python (Version 3.11.14),^24^ models were fit using the mgcv R package,^29^ and all statistical analysis was performed using R (Version 4.6.0),^25^ with α = .20 used for feature selection and α = .05 for all other hypothesis testing.

### Pitch-level variables

Statcast data provides pitch velocity, pitch type, spin rate, arm angle, and measured vertical and horizontal movement for each pitch, among other features.

Statcast defines arm angle as the angle with respect to the horizontal of a line connecting the shoulder and the hand at ball release.^1^ A pitcher who throws the ball with their arm parallel to the horizontal (e.g. standing upright, shoulder at 90° relative to trunk, elbow straight) has a Statcast arm angle of 0°. This measure is related to, but not synonymous with, arm slot as it is defined in the biomechanical literature. Pitchers with the same arm slot may have different Statcast arm angles depending on their trunk tilt at ball release, for example.

We classify both the slider (SL) and sweeper (ST), a new slider sub-type introduced by Statcast in 2023,^14^ as sliders. Sinkers (SI), four-seam fastballs (FF), and cut fastballs or “cutters” (FC) were categorized as fastballs.^12^ In our analysis, we considered a pitcher’s mean arm angle as well as their velocity, spin rate, and usage for “fastballs” (FF, SI, and FC) and “sliders” (SL + ST), averaged over pitches thrown by the pitcher during that season.

### Types of injury

We employed a public database of pitchers’ UCLR and transaction data from the MLB Stats API to identify pitcher injuries.^13,26^ For injuries not resulting in UCL reconstruction, transaction data was parsed using regular expressions (regex) to identify IL placements by matching case-insensitively to keywords. Any identified cases of UCLR were tagged as such, while identifying injury type from transaction data was more nuanced.

When parsing transaction data for injury type, descriptions corresponding to acute trauma were proactively excluded on the grounds that they were of indeterminate relevance to pitching (e.g. abrasions, bruises, fractures) or not pitching-related (e.g. collision, hit by pitch, etc.). Any IL placements containing these terms were excluded: fracture, contusion, laceration, hematoma, abrasion, bruise, avulsion, puncture, collision, hit by pitch, hbp.

Remaining descriptions were matched to either “elbow” or “forearm” based on their contents. Descriptions containing any of “elbow,” “ulnar,” “UCL,” “Tommy John”, or “epicondyl” (to match any of epicondyle, epicondylitis, etc.) were assigned to “elbow.” Any entries not already categorized as an elbow injury were assigned to “forearm” if they contained “forearm,” “pronator,” or “flexor” (only when not preceded by hip or elbow).

The groups of UCLR, Elbow, Forearm, and their union (“Elbow/Forearm”) were studied for injury risk factors. Seasons were categorized by whether they contained a UCLR procedure and/or IL placement of the corresponding type, not constrained to be mutually exclusive, meaning a pitcher who sustained both an elbow and forearm injury could appear in both.

### Inclusion criteria

#### Position players pitching

Because Statcast data logs all pitches thrown in any MLB game, it occasionally includes non-pitchers. MLB teams are permitted, under limited circumstances, to use position players in lieu of an actual pitcher. To address this, pitchers were required to have faced at least 45 batters and have at least one fastball (FF, SI, or FC) tracked by Statcast.

#### Prior season workload

In addition to analyzing the set of all seasons meeting these criteria, we also report two sub-analyses of pitchers with 30 IP the prior season. All three analyses used the same methodology, save for the additional criterion and the timing of injury. One sub-analysis predicted same-season injury risk (e.g. 2021 data to predict incidence of injury in 2021) and the other predicted injury risk the next year (e.g. 2021 data to predict injury in 2022).

### Feature selection and GAM fit

Candidate features for each fit were evaluated using a bivariate screen against the corresponding injury type, and α = .20 was used for inclusion. Selected features were included in a generalized additive model (GAM) fit with binomial response for each variable of interest using a reduced maximum likelihood (REML) optimization method.

Average pitcher arm angle and velocity, spin rate, and usage for fastballs and sliders were evaluated for inclusion. For both fastballs and sliders, main effects for arm angle, velocity, usage, and spin rate were included as spline smooths, where up to 3 degrees of freedom could describe a non-linear relationship to injury risk. Tensor product smooths (*ti* in mgcv) representing the marginal interactions of arm angle with velocity and usage were also included for both pitch types, as were the marginal interaction between usage and velocity. Additionally, because of prior findings^19^ about the relationship between fastball velocity and slider usage, we also included that interaction across pitch types.

All smoothed terms were fit using cubic splines with shrinkage (*bs* = ′*cs*′) and basis dimension of 4 (*k* = 4) for smooth terms. This provides the model with spline terms which nominally have three degrees of freedom and the ability to penalize both the curvature and contribution of the term to zero. In practice, together with REML optimization, this allows additional feature selection by the model when fitting and greatly reduces the effective degrees of freedom for each term and model.

We report effective degrees of freedom (*n_dof_*) for each term and fit, as well as the nominal degrees of freedom for each fit. For main effect terms, average marginal effect (AME) odds ratios are reported to summarize average effect. Regression p-values and Nagelkerke pseudo-*R*^2^ are presented for each fit, along with feature-level p-values and risk profile plots.

## Results

Of 5,335 candidate pitcher-seasons, N = 3,812 were MLB pitchers who faced at least 45 hitters in that season. The included pitcher-seasons had a 11.1% incidence of any qualifying elbow or forearm injury (*n_pos_* = 424). We found 2.1% incidence of UCL reconstruction (*n_pos_* = 80), 8.1% incidence of elbow injury (*n_pos_* = 307), 3.9% incidence of forearm injury (*n_pos_* = 149), and 0.8% incidence (*n_pos_* = 32) of forearm and elbow injury in the same season. Identified injury reasons for each group are reported in *Table 1*. UCLR entries which reference the forearm (e.g. forearm strain) reflect IL transaction reasons given for a pitcher who underwent UCLR during that season, indicating the overlap between forearm and elbow injury reasons from MLB IL data. Of the 80 pitcher-seasons in the UCLR group, 23 (29%) had only forearm-related IL reasons, while 2 had a forearm IL stint followed by an elbow IL stint. 8 pitchers who underwent UCLR were missing a measured slider velocity and dropped when restricting to complete cases prior to modeling, leaving *n_pos_* = 72 for UCLR. Time from first forearm IL placement in a season to UCLR ranged from [9, 177] days with *t_mean_* = 44, *t_median_* = 27.

**Table 1:** Top 10 IL placement reasons by injury group for MLB pitchers from 2020 to 2025. Elbow placement group is inclusive of UCLR group; references to “forearm” in UCLR group indicate cases where a pitcher undergoing Tommy John surgery were on the IL for a reason citing their forearm, e.g. “forearm strain.”

| Rank | UCL Reconstruction | n | Elbow | n | Forearm | n |
| --- | --- | --- | --- | --- | --- | --- |
| 1 | Elbow inflammation | 16 | Elbow inflammation | 114 | Forearm strain | 53 |
| 2 | Elbow sprain | 10 | Elbow strain | 26 | Forearm inflammation | 21 |
| 3 | Forearm strain | 9 | Elbow sprain | 25 | Flexor strain | 15 |
| 4 | Elbow strain | 6 | Elbow discomfort | 15 | Forearm tightness | 13 |
| 5 | Forearm inflammation | 5 | Elbow flexor strain | 12 | Forearm flexor strain | 7 |
| 6 | UCL sprain | 4 | Forearm strain | 9 | Forearm tendinitis | 5 |
| 7 | Elbow discomfort | 4 | Elbow tendinitis | 7 | Forearm extensor strain | 4 |
| 8 | Strained elbow | 3 | Elbow soreness | 7 | Forearm soreness | 4 |
| 9 | Forearm tightness | 3 | UCL sprain | 6 | Flexor muscle strain | 3 |
| 10 | Elbow injury | 2 | Forearm inflammation | 5 | Flexor pronator strain | 3 |

**Table 2:** Feature-level p-values for bivariate screen by outcome variable. Bold indicates features selected for final fits (p < .20); asterisks indicate statistical significance (* < .05, ** < .01, *** < .001).

| Feature | UCLR All Pit. | Elbow All Pit. | Forearm All Pit. | Elbow or Forearm All Pit. | UCLR Min. 30 IP Same Year | Elbow Min. 30 IP Same Year | Forearm Min. 30 IP Same Year | Elbow or Forearm Min. 30 IP Same Year | UCLR Min. 30 IP Next Year | Elbow Min. 30 IP Next Year | Forearm Min. 30 IP Next Year | Elbow or Forearm Min. 30 IP Next Year |
| --- | --- | --- | --- | --- | --- | --- | --- | --- | --- | --- | --- | --- |
| Arm Angle | 0.385 | **0.009\*\*** | **0.004\*\*** | **0.001\*\*** | 0.321 | **0.071** | **0.004\*\*** | **0.001\*\*** | 0.737 | **0.060** | **0.039\*** | **0.005\*\*** |
| Fastball Velocity | 0.556 | **0.158** | **0.052** | **0.015\*** | 0.484 | 0.826 | 0.217 | 0.298 | 0.738 | 0.419 | 0.368 | **0.127** |
| Fastball Usage | **0.056** | **0.018\*** | **0.072** | **0.026\*** | 0.330 | **0.192** | **0.150** | 0.209 | 0.246 | 0.334 | **0.096** | 0.210 |
| Fastball Spin Rate | **0.063** | 0.204 | **0.009\*\*** | **0.027\*** | 0.269 | 0.594 | 0.204 | 0.275 | **0.064** | 0.221 | 0.242 | 0.222 |
| Slider Velocity | 0.256 | **0.019\*** | **0.0006\*\*\*** | **0.00009\*\*\*** | 0.406 | 0.206 | **0.043\*** | **0.024\*** | 0.712 | 0.231 | **0.057** | **0.031\*** |
| Slider Usage | **0.052** | **0.016\*** | 0.618 | **0.024\*** | **0.066** | **0.042\*** | 0.902 | **0.114** | **0.144** | **0.096** | 0.960 | **0.159** |
| Slider Spin Rate | **0.026\*** | **0.182** | **0.0003\*\*\*** | **0.003\*\*** | 0.258 | 0.870 | **0.021\*** | **0.150** | **0.179** | 0.611 | **0.069** | **0.165** |
| Fastball Velocity × Arm Angle | 0.822 | 0.335 | **0.067** | **0.062** | 0.768 | 0.372 | 0.214 | 0.314 | 0.679 | 0.692 | 0.324 | 0.762 |
| Fastball Usage × Arm Angle | 0.359 | **0.044\*** | 0.334 | **0.012\*** | 0.301 | **0.023\*** | 0.342 | **0.006\*\*** | **0.132** | **0.097** | 0.317 | **0.058** |
| Slider Velocity × Arm Angle | 0.873 | 0.486 | **0.185** | **0.190** | 0.951 | 0.595 | 0.334 | 0.255 | 0.956 | 0.833 | 0.617 | 0.545 |
| Slider Usage × Arm Angle | 0.767 | **0.045\*** | **0.136** | 0.288 | 0.949 | 0.618 | 0.401 | 0.798 | 0.356 | 0.312 | 0.509 | 0.944 |
| Fastball Usage × Fastball Velocity | 0.224 | 0.827 | 0.224 | 0.587 | 0.203 | 0.491 | 0.816 | 0.435 | 0.833 | 0.941 | 0.825 | 0.991 |
| Slider Usage × Slider Velocity | 0.905 | 0.821 | **0.006\*\*** | **0.083** | 0.986 | 0.980 | 0.359 | 0.500 | 0.608 | 0.472 | 0.527 | 0.350 |
| Slider Usage × Fastball Velocity | 0.868 | **0.191** | 0.807 | **0.119** | 0.807 | 0.673 | 0.978 | 0.682 | 0.313 | 0.417 | 0.915 | 0.294 |

**Table 3:**
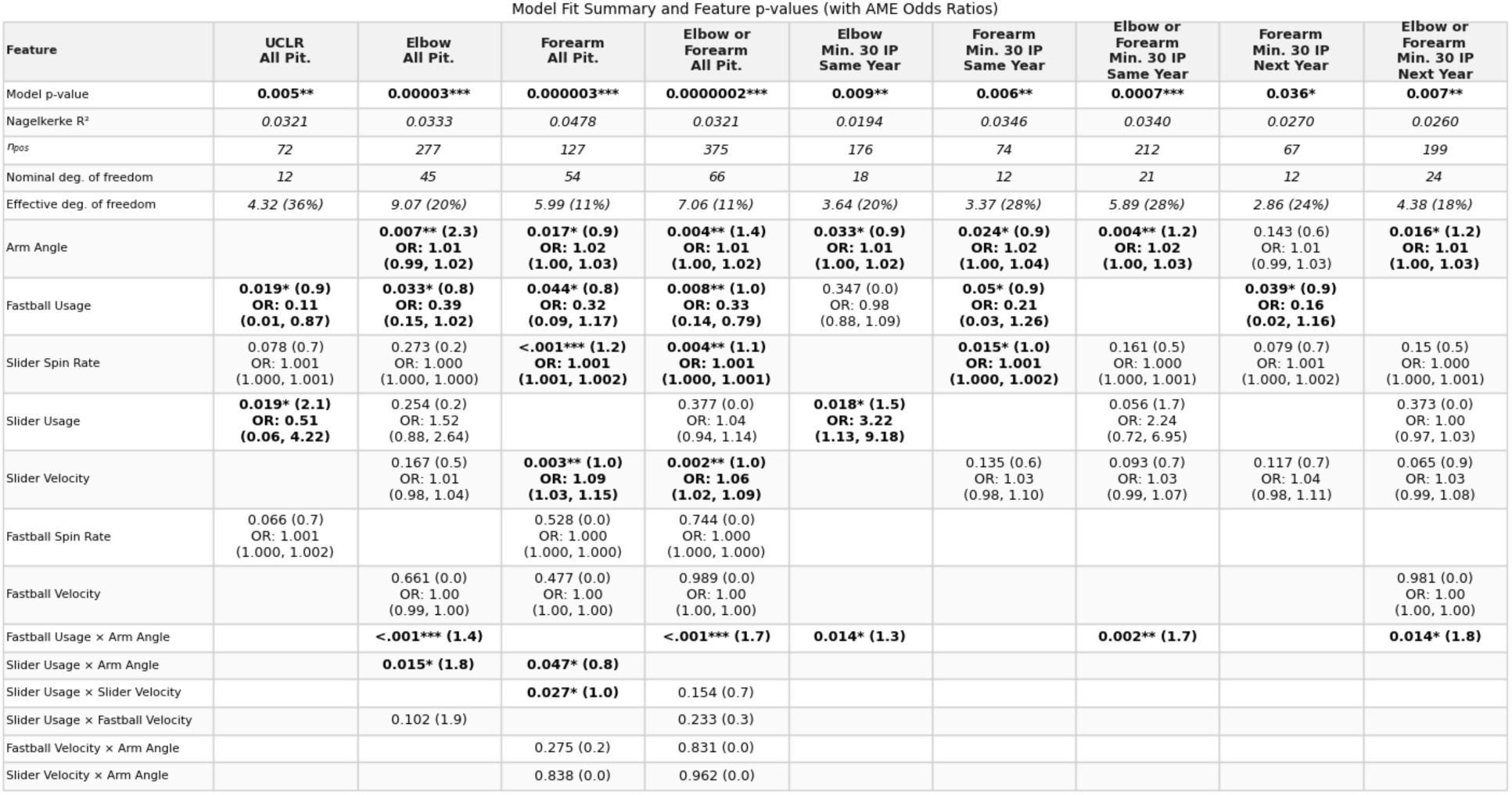
Fit summary information and feature-level p-values, degrees of freedom, and average marginal effect odds ratios shown. Variables show p-values, with n_dof_ in parentheses, and AME odds ratios with 95% confidence interval are included for main effects. Same-season UCLR (p = .27), next-season UCLR (p = .081), and next-season Elbow (p = .054) were not significant and omitted for brevity. (* < .05, ** < .01, *** < .001).

### Feature selection

Arm angle, slider usage, and slider spin rate were the most frequently selected terms in the bivariate screen, with 9 fits selecting them. Slider velocity, fastball usage, and the marginal interaction between fastball usage and arm angle were selected in 7 fits. Fastball velocity alone was only selected in 4 fits.

### Generalized additive model fits

Arm angle was significant in 7 of the 9 statistically significant fits, the most of any feature, and it was positively associated with the incidence of injury across all 7. In the Elbow injury fit across All Pitchers, modeled injury incidence peaked near 45° before decreasing; in the other 6 fits, injury incidence monotonically increased as a function of arm angle.

Average marginal effect, computed as the average effect over the range of the variable of interest, provides a point estimate of a potentially non-linear relationship. Odds ratios derived from AME for arm angle ranged from [1.005, 1.022], with the lowest value corresponding to the Elbow, All Pitchers fit. The non-linearity of risk as a function of arm angle means that the AME-derived odds ratio overestimates the effect at arm angle and underestimates it at high arm angle.

In the 6 fits in which it was statistically significant, fastball usage was associated with monotonically decreasing injury incidence. AME odds ratios for fastball usage ranged from [0.11, 0.38]. The main effects of arm angle and fastball usage were the two most frequently significant features, followed by their marginal interaction, which was significant in 5 fits.

While the bivariate relationships between arm angle and fastball usage were consistent across all fits, their marginal effects exhibited considerable heterogeneity relative to main effects. At high arm angle, increased fastball usage was no longer protective, and injury remained elevated. In the low arm angle regime, the effect of fastball usage dominated, and low fastball usage was associated with elevated risk, muting any protective effect of throwing from a low arm angle.

**Figure 1:**
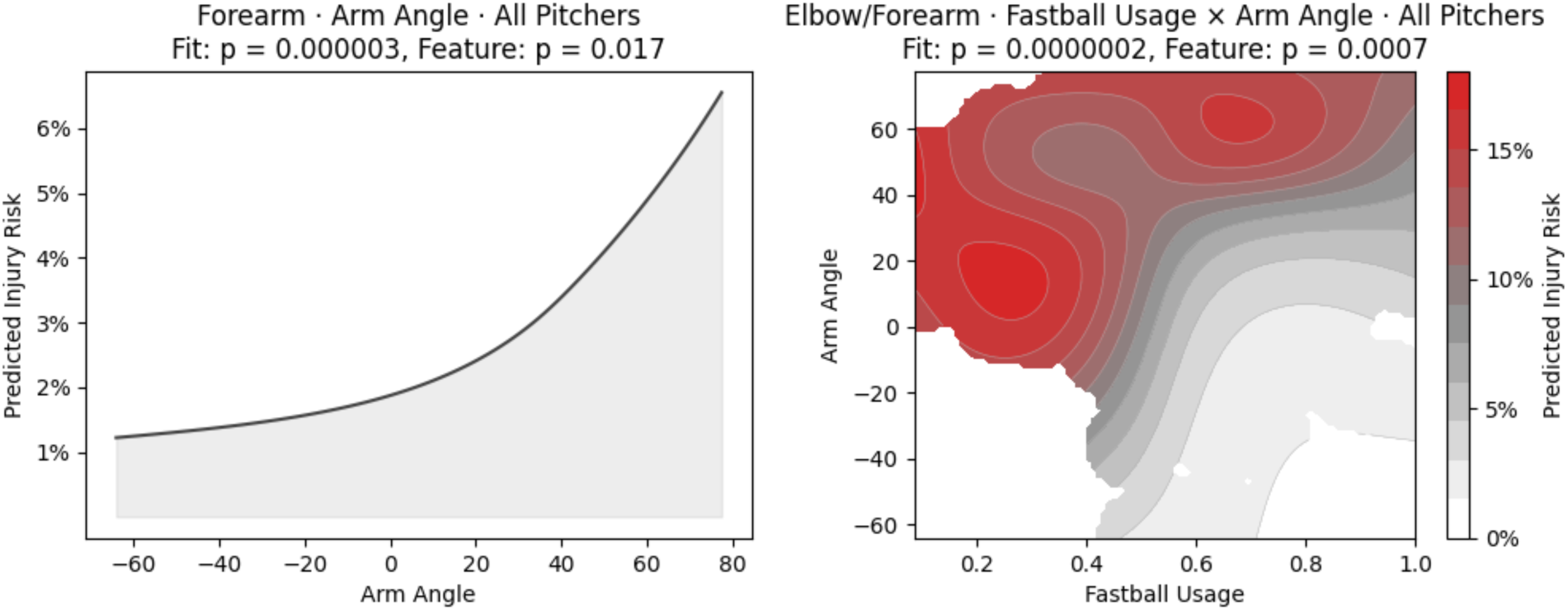
Modeled probability of (a) forearm injury as a function of arm angle and (b) elbow/forearm injury as a function of fastball usage and arm angle, holding other inputs constant at cohort medians. (a) Predicted forearm injury incidence was a monotonically increasing, concave-up function of arm angle. (b) At elevated arm angles, any protective effect of increasing fastball usage is greatly diminished, while at low arm angles, modeled elbow/forearm injury incidence largely depends on fastball usage.

Slider spin rate and velocity were also significantly associated with incidence of injury. Across all pitchers, slider velocity predicted incidence of Forearm injury (*p* = .003, *AME OR* 1.088, 95% *CI* [1.026, 1.155]) and Elbow/Forearm injury (*p* = .002, *OR* 1.055, 95% *CI* [1.018, 1.093]). Spin rate also significantly predicted injury incidence among all pitchers for Forearm (*p* = .0003, *AME OR* 1.0013, 95% *CI* [1.0006, 1.0021]) and Elbow/Forearm (*p* = .004, *AME OR* 1.0006, 95% *CI* [1.0002, 1.0011]), as well as same-season Forearm injury among pitchers who threw at least 30 IP the prior season (*p* = .015, *OR* 1.0010, 95% *CI* [1.0001, 1.0019]). Median slider spin rate was 2,429 rpm, so a 1.0013 AME odds ratio indicates that a 100 rpm increase in slider spin rate would coincide with a 14% increase in the risk of Forearm injury on average.

The effect of slider usage was more mixed across fits. In the two fits where slider usage significantly predicted injury incidence, average marginal effect odds ratios were 0.51 (UCLR, All Pitchers) and 3.22 (Same-season Elbow, 30 IP). In general, predicted risk increased at low slider usage before peaking and eventually decreasing at high slider usage, but precise end behavior varied by fit, including in the two where this feature was significant.

The marginal interaction effect of slider usage with slider velocity was significantly (*p* = .027) associated with Forearm injury among all pitchers, where slider usage moderated the effect of velocity by increasing the modeled risk as a function of velocity at higher slider usage. The marginal effect of slider usage interacted with arm angle significantly predicted incidence of Elbow injury among all pitchers (*p* = .015), with the highest risk coming at elevated fastball usage and an arm angle between 40° and 60°. pseudo-*R*^2^ values ranged from .019 to .048, and effective degrees of freedom for significant fits ranged from 2.86 to 9.07. 8 of the 12 fits performed were significant at *p* < .01, and one additional fit was significant at *p* < .05. Despite selecting many features in some fits, including interaction features with many nominal degrees of freedom, the GAM fits using REML effectively limited their contribution by shrinking many coefficients to 0, including fastball velocity, which was regularized out of all fits for which it was selected.

**Figure 2:**
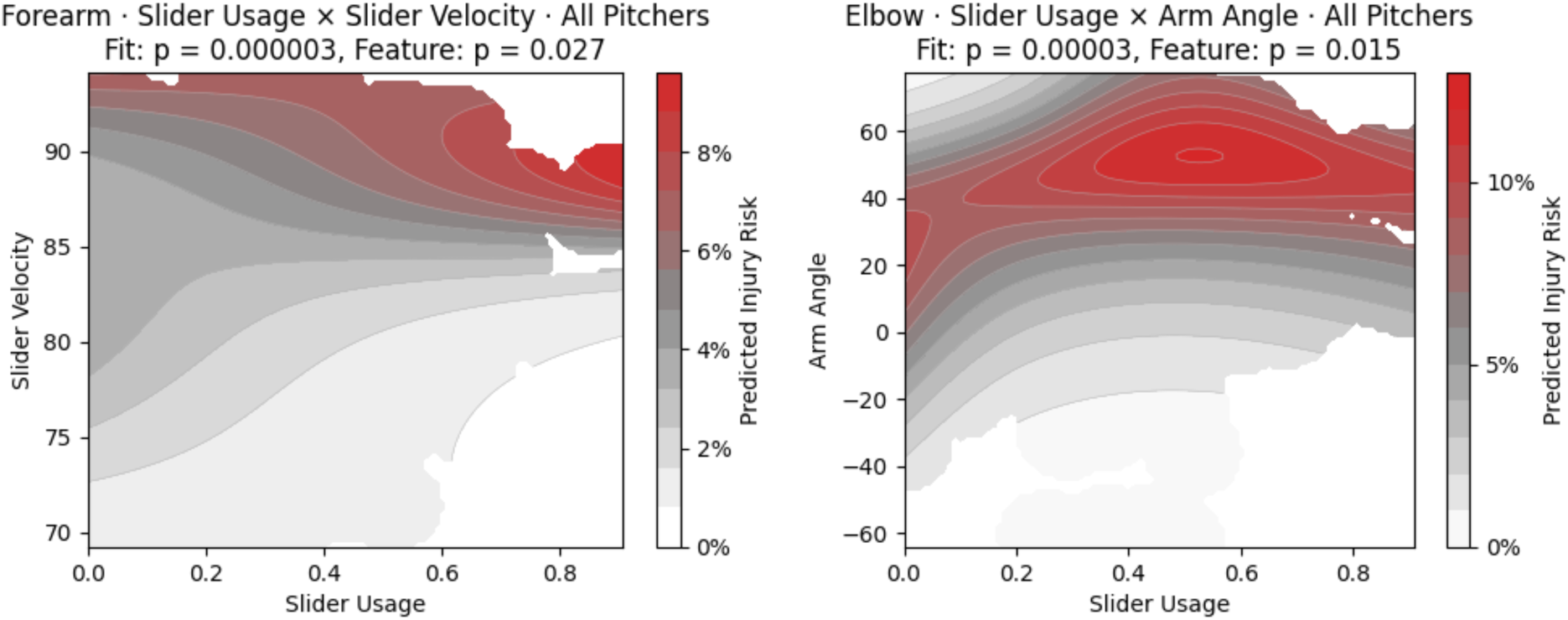
Modeled probability of (a) forearm injury as a function of slider usage and velocity and (b) elbow injury as a function of fastball slider and arm angle, holding other inputs constant at cohort medians. (a) Highest risk of forearm injury occurs at elevated velocity and usage. (b) Increasing slider usage has little effect on predicted risk of elbow injury at low arm angle, but at high arm angles is associated with rapidly increasing risk.

## Discussion

We found that arm angle, fastball usage, and their interaction effect, not fastball velocity, significantly predicted injury risk among MLB pitchers. Across the nine significant fits presented here, one of these three features was significant in each one. Of the 3 features, an average of 2 were significant per fit. The predictive power of arm angle and fastball usage persisted when predicting injury risk the following season. The relationship between fastball velocity and injury risk shown elsewhere in the literature was observed in bivariate feature screens, both for main effects and the interaction with arm angle. However, built-in regularization selected the interaction arm angle, fastball usage, and/or their interaction in lieu of velocity each time when fitting full models.

The physiological basis for the arm angle finding is not straightforward. Laboratory studies of arm slot and elbow torque disagree on direction: Aguinaldo and Chambers found sidearm delivery produced higher valgus torque than an overhand slot,^2^ while a more recent comparison across skill levels found the opposite relationship in professional pitchers and no clear relationship in high school pitchers.^15^ Statcast arm angle is also a composite of shoulder abduction and trunk tilt at release rather than a controlled biomechanical arm slot measurement, and it reflects cumulative, season-long exposure rather than peak torque on a single pitch. Both distinctions may help explain why the direction of the arm angle effect observed here does not map cleanly onto single-pitch torque studies.

The protective association of fastball usage is similarly not explained by single-pitch torque. Fastballs generate the highest peak and cumulative elbow varus torque of any commonly thrown pitch, in both professional and high school pitchers.^4^ Werner et al. found that triceps, flexor-pronator, and anconeus activity increase near peak valgus stress, suggesting these muscles act as dynamic stabilizers of the UCL.^28^ A pitcher relying on a consistent, well-conditioned fastball delivery across a season may recruit this stabilizing musculature more reliably than one relying more heavily on breaking balls, where altered forearm pronation and supination demands could reduce the same stabilization. This is consistent with our finding that slider velocity and spin rate, not fastball velocity, predicted forearm and elbow/forearm injury. Testing the muscular mechanism directly would require electromyographic or wearable-sensor data alongside Statcast-derived usage and velocity.

Prior work has identified fastball velocity as a predictor of injury risk across seasons,^16^ but our results suggest that arm angle and usage may be a more important predictor of medial elbow injury than fastball velocity, a relationship heretofore underexplored given the recent addition of arm angle data to public Statcast data.

Unlike fastballs, slider velocity and spin rate were significant predictors of injury risk in both Forearm and Elbow/Forearm fits, consistent with prior results.^16,22^ Additionally, a significant interaction was observed between slider usage and arm angle when predicting Elbow injury, where increasing slider usage carried little additional risk at low arm angles but greatly increased risk at higher ones. The link between arm angle and slider usage was weaker than the link to fastball usage, but that relationship may still warrant further study, given the predictive power of arm angle, slider usage, velocity, and spin rate individually.

The relationship between slider usage and slider velocity appears to be consistent with the interaction effect observed by Oeding et al. (2024) between slider usage and fastball velocity.^19^ The authors in that work, using SHAP (SHapley Additive exPlanations) to model feature contributions to their decision tree model’s output, observe little effect on predicted risk for slider usage and fastball velocity at all but high slider usage. However, at high slider usage, SHAP values indicate a strong, positive relationship between slider usage and fastball velocity. We found that the relationship between slider usage and fastball velocity was not significant, but the relationship between slider usage and slider velocity was significant. This suggests that the model created by Oeding et al. (2024) may have identified an injury risk posed by throwing a lot of sliders at high velocities, using fastball velocity as a proxy.

Existing analyses are mixed in the reporting of *R*^2^ for predicting elbow injury. However, Chalmers et al. (2016) do report Nagelkerke pseudo-*R*^2^ values to quantify the strength of pitch velocity and other features as predictors of UCLR. They find that peak pitch velocity explains 3.6% of the variance in UCLR among MLB pitchers (*R*^2^ = .036). Additional pitch-level data explain an additional 1.3% of the variance (*R*^2^ = .049), and the combined fit when including age and BMI gives *R*^2^ = .068.^3^ These benchmarks suggest fit-level *R*^2^ values from .019 to .048, including .032 for UCLR, using only pitch-level data are consistent with the best available estimates.

Our analysis is not without limitations. Notably, we studied a broad set of injury descriptions from MLB Injured List placements, which posed challenges when assessing injury severity or disentangling elbow from forearm. The numerous instances of players who underwent UCL reconstruction while on the IL for a forearm injury, 29% of UCLR cases, illustrated that interrelationship. This finding supports the decision to consider forearm and elbow injuries together and is consistent with prior findings about the link between forearm injury and UCL injury.^5,30^ Combining them was an a priori anatomical decision as much as an empirical one, since the flexor-pronator mass and the anterior bundle of the UCL form a continuous medial elbow complex that shares a valgus load path.^6,21^ The tradeoff is that the combined outcome cannot separate a primary muscular or tendinous injury from a primary ligamentous one. IL reasons are the information MLB teams choose to disseminate, often at the onset of player injury, not a medical diagnosis, meaning they may be a noisy proxy.

In addition to IL reasons complicating the assessment of injury severity, measuring days lost to injury from MLB transaction data alone is a prohibitively difficult task, because such an estimate using transaction data requires numerous medical and strategic assumptions. In keeping with Oeding et al.’s approach to use IL placements as an outcome,^19^ we used publicly available data to study injury incidence. However, a dataset like MLB’s Health and Injury Tracking System (HITS) offers a high-quality measure of days lost by looking at medical clearance date instead.^10,27^ Future work should use days lost to injury and injury history from datasets like HITS, along with other demographic and pitch-level variables, to quantify the relationship between injury severity and the variables identified here.

Furthermore, while we find that arm angle and fastball usage are a stronger signal than fastball velocity for predicting incidence of elbow and forearm injury among MLB pitchers, it is important to properly contextualize that result. This relationship was observed among MLB pitchers with median fastball velocity of 93.7 mph across four-seam, sinkers, and cutters. Our results are not inconsistent with velocity being an injury predictor on its own, nor do they necessarily generalize to lower levels of competition with lower, more heterogeneous fastball velocities. The results presented here merely suggest that the interaction of arm angle and fastball usage is a better indicator of injury risk than fastball velocity among MLB pitchers. This supports the findings of Keller et al. (2016) that fastball usage, not velocity, predicts injury risk,^7^ and it suggests that arm angle is responsible in part for moderating that risk.

The significant association between pitch-level characteristics and arm angle as predictors of injury incidence underscores the importance of arm angle data in understanding MLB pitcher injuries. Future work should investigate the relationship between arm angle and pitch selection for other pitch types. For example, leading surgeons like Dr. Keith Meister have hypothesized a link between arm injuries and sweeper usage.^8^ Future work should investigate further whether a relationship exists between arm angle, sweeper usage, and injury. For example, studying the relationship between horizontal break, arm angle, and injury for sliders would allow an assessment of sweeper risk as a function of arm angle. Similar approaches can be applied to other pitch types to understand whether they also exhibit a non-linear relationship between arm angle, usage, and injury.

## Conclusion

Arm angle, fastball usage, and their interaction, not fastball velocity, significantly predicted incidence of elbow injury among MLB pitchers from 2020 to 2025. Despite increasing fastball usage being broadly associated with lower incidence of injury, this effect was moderated at high arm angles, where increasing fastball usage was no longer associated with lower injury risk. The interaction of arm angle and fastball usage remains a significant predictor of elbow or forearm injury across seasons over and above fastball velocity, which was not significant in any fit. Pseudo-*R*^2^ values using arm angle, usage, and velocity for fastballs and sliders range from .019 to .052 among the fits presented, in line with existing benchmarks for pitch tracking-based predictions of elbow injury among MLB pitchers.

## Data Availability

All input data for the present study are freely available at the sources listed. Engineered data used for analysis may be made available upon reasonable request to the authors.

https://docs.google.com/spreadsheets/d/1gQujXQQGOVNaiuwSN680Hq-FDVsCwvN-3AazykOBON0

https://baseballsavant.mlb.com

https://statsapi.mlb.com

